# Associations of Blood Inflammatory Cytokines and Hair Cortisol with Depression Symptoms and Cognitive Functioning in Huntington’s Disease

**DOI:** 10.1101/2023.07.09.23292433

**Authors:** Hiba Bilal, Stuart J. McDonald, Julie C. Stout, Ian H. Harding

## Abstract

**Background:** Huntington’s disease (HD) is an inherited neurodegenerative disease involving progressive motor abnormalities, cognitive decline, and psychiatric disturbances. Depression and cognitive difficulties are among the most impactful symptoms of HD, yet the pathogenesis of these symptoms is not fully understood. HD involves low-level chronic inflammation and dysregulation of the hypothalamic-pituitary-adrenal (HPA) axis, which are linked to depression and cognitive impairment in non-HD populations. However, previous research on the relationships of these pathologies with depression and cognition in HD is limited and inconsistent.

**Methods:** Fifty-three adults with the HD gene expansion (30 pre-manifest, 23 manifest) completed measures of depression and cognitive functioning. Forty-eight out of 53 participants provided hair samples for quantification of cortisol, and 34 participants provided blood samples for quantification of peripheral inflammatory cytokines. We examined the associations of four cytokines (interleukin (IL)-6, IL-10, IL-1β, and tumour necrosis factor (TNF)-α) and cortisol levels with depression and cognitive scores.

**Results:** In unadjusted models, higher levels of plasma IL-6, IL-10, and TNF-α correlated with higher depression scores, and higher levels of IL-10 and TNF-α correlated with poorer cognitive performance. After controlling for age, sex, and body mass index, only the correlations of IL-10 with depression and cognitive performance remained significant. No correlations were evident with hair cortisol.

**Conclusions:** Peripheral inflammation is associated with depression symptoms and cognitive impairment in HD. Our findings suggest that interactions between the immune and nervous systems are important in HD, and that inflammatory cytokines may be suitable as therapeutic targets for future clinical trials in HD.

## Introduction

Huntington’s disease (HD) is an inherited neurodegenerative disease caused by an abnormal expansion in the number of cytosine-adenine-guanine (CAG) repeats in the huntingtin gene, which causes a mutated form of the huntingtin protein to be encoded and expressed throughout the body [1]. Motor abnormalities are traditionally considered to be the hallmark feature of HD, and clinical diagnosis of HD requires the unequivocal presence of motor signs [2]. Yet, a growing body of research indicates that cognitive and psychiatric disturbances have a more significant impact on the lives of people with HD than motor symptoms [3–5]. Moreover, psychiatric disturbances and subtle changes in cognitive functioning can emerge 10 to 15 years before clinical diagnosis of HD [6–8] and may consequently begin to affect quality of life early in the disease course. Therefore, identifying and addressing the factors that contribute to cognitive and psychiatric difficulties in HD is essential for optimising the quality of life of HD patients.

Depression is one of the most common psychiatric syndromes to affect HD CAG expansion carriers [9, 10]. Depression can emerge in any stage of HD but appears most commonly just before HD diagnosis and in the early stages of HD [11, 12]. Moreover, depression does not appear to increase in prevalence or severity as HD progresses, instead following a non-linear course across the disease [11–14]. The aetiology of depression in HD appears to involve both neurobiological and psychosocial factors [15] but is not well understood. Unlike depression, cognitive functioning is known to decline as HD progresses, along with other disease symptoms like motor functioning and apathy [7, 16]. Cognitive deficits in HD are thought to result from progressive brain atrophy caused by the expression of mutant huntingtin throughout the body [17]. However, other factors are also likely to affect cognitive functioning in HD, given that the onset and progression of cognitive decline in HD is highly variable, and because cognitive deficits do not appear to map onto brain changes in pre-manifest HD [17–19].

In non-HD populations, depression and cognitive impairment are strongly linked to dysregulation of the hypothalamic-pituitary-adrenal (HPA) axis, and inflammation in the central nervous system (CNS) and periphery [20–27]. Notably, both the HPA axis and the immune system are known to be dysregulated in HD. Specifically, people in the pre-manifest stage of HD exhibit elevated HPA axis activity, indicated by higher salivary and plasma cortisol levels relative to healthy controls and manifest HD participants; in contrast, people with manifest HD exhibit a hypoactive HPA axis, evidenced by lower salivary cortisol levels relative to healthy controls and participants with pre-manifest HD [28–31]. HD CAG expansion carriers also exhibit low-level chronic inflammation in the CNS and periphery, evidenced by increased microglial activation within the striatum and cortex [32–35] and higher levels of inflammatory cytokines within brain tissue, cerebrospinal fluid, and blood plasma relative to control groups [36–40]. Low-level inflammation is evident years before the clinical diagnosis of HD, and many inflammatory markers have been shown to increase across disease stages or correlate with measures of disease progression in HD [33, 34, 36].

Current evidence for the relationships of HPA axis functioning with depression and cognition in HD is inconsistent. Several studies have reported that cortisol levels are associated with depression symptoms and cognitive functioning in HD [29, 30, 41], whereas other studies have reported no significant associations among these measures [42, 43]. Moreover, the specific protocols and indices used to measure HPA axis activity also differ considerably across studies, which further limits comparisons among previous findings. Meanwhile, the relationship of inflammation with depression and cognition in HD has only been investigated in one study [44], in which peripheral inflammatory cytokines were significantly associated with cognitive performance, but not depression symptoms. Given the limited research in this area, and discrepancies among existing studies, the relationships of HPA axis functioning and inflammation with depression and cognition in HD remain poorly understood.

In this study, we assessed HPA axis functioning by quantifying cortisol levels in hair samples. Hair samples allow for quantification of cumulative cortisol release across several months, which render them suitable for measuring *chronic* HPA axis functioning relative to traditional cortisol measures (i.e., blood, urine, and saliva) that are influenced by daily diurnal rhythms and other sources of contextual variability [45]. Numerous studies have assessed hair cortisol in relation to depression and cognitive functioning in neurologically healthy groups [46–49], whereas only one study has assessed hair cortisol in pre-manifest HD [43]. We also measured systemic or peripheral inflammation by quantifying four inflammatory cytokines (interleukin (IL)-6, IL-10, IL-1β, and tumour necrosis factor (TNF)-α) in blood plasma, using highly sensitive immunoassays. These cytokines are known to be elevated in HD relative to healthy control groups [36, 38], and have previously been linked to depression and cognitive functioning in non-HD populations [21, 50]. The primary aims of this study were to examine the associations of chronic HPA axis activity (hair cortisol concentrations) and systemic inflammation (blood-based inflammatory cytokines) with depression symptoms and cognitive performance in pre-manifest and early manifest HD. A secondary aim was to examine whether these associations differed according to disease stage (pre-manifest versus manifest HD groups). We also explored the associations of chronic HPA axis activity and systemic inflammation with demographic and clinical outcomes in HD.

## Method

### Participants

Fifty-three people with the HD CAG expansion (CAG repeat length ≥ 36) were recruited through the Experimental Neuropsychology Research Unit and Clinical Cognitive Neuroscience (ENRU-CCN) Laboratory Research Participant Registry, HD specialist clinics and advocacy groups in Australia, and social media. We recruited people in the pre-manifest or early manifest stages of HD. The pre-manifest group (*n* = 30) consisted of CAG-expanded participants who did not meet criteria for diagnosis (based on motor symptom expression) at the time of participation. Two pre-manifest participants had a CAG repeat length of 38, which indicates reduced penetrance of the disease phenotype [51]. The early manifest group (*n* = 23) included clinically diagnosed participants who were in Stage I or II of HD (Total Functional Capacity [TFC] Score ≥ 7) [52]. The TFC scale assesses the ability to independently carry out activities such as domestic chores, management of finances, personal grooming, and hygiene. Total scores can range from 0 to 13, with a lower score indicating poorer functional capacity and greater disease severity. We also calculated a Disease Burden Score (DBS; (CAG – 35.5) × Age) for each participant to estimate lifelong exposure to mutant huntingtin [53].

Exclusion criteria for this study included psychiatric conditions other than depression and anxiety, neurological disorders other than Huntington’s disease, previous significant (i.e., moderate to severe) traumatic brain injury and/or concussion sustained in the past 12 months, consumption of excessive alcohol (>10 standard drinks per week) or illicit drugs, and current participation in a clinical drug trial. Additional exclusion criteria for the blood collection phase were autoimmune or inflammatory disorders, and current use of anti-inflammatory or immunomodulatory medications. For the hair sample collection phase, individuals with hair length <3 cm were excluded.

From the 53 participants recruited, 48 provided hair samples, and 34 provided blood samples. Several participants were unable to provide a blood sample because of restrictions, either due to the COVID-19 pandemic, or because no collection centre was available within a reasonable distance from their residence. Chi-square tests and independent-samples *t*-tests (or Mann-Whitney *U* tests for non-normally distributed variables) indicated that the group of participants who provided a blood sample included significantly more people with a prior diagnosis of major depressive disorder, compared to those who did not provide a blood sample (χ^2^(1) = 7.30, *p* = .007), even though depression scores did not significantly differ between blood sample providers and non-providers. No significant differences were evident for any demographic variables, disease severity outcomes, or cognition scores (all *p* values > .05). With respect to hair samples, of the 53 total participants, four had insufficient hair for sample collection or quantification, and one participant opted out. Therefore, statistical analyses were conducted using data from 48 hair samples (26 pre-manifest, 22 manifest) and 34 blood samples (20 pre-manifest, 14 manifest).

Table 1 reports descriptive statistics for the hair and blood sample subgroups, stratified by disease stage (pre-manifest and manifest groups). In both the hair and blood sample subgroups, participants in the manifest group were significantly older, and had poorer functional capacity and cognitive performance relative to the pre-manifest group. In the hair sample subgroup only, the manifest group also had significantly greater disease burden than the pre-manifest group; this between-groups difference approached significance (*p* = .07) in the blood sample subgroup. Depression scores did not significantly differ between pre-manifest and manifest participants in either subgroup; however, among hair sample providers, significantly more manifest participants were using antidepressants relative to pre-manifest participants.

## Descriptive Statistics

**Table 1.**
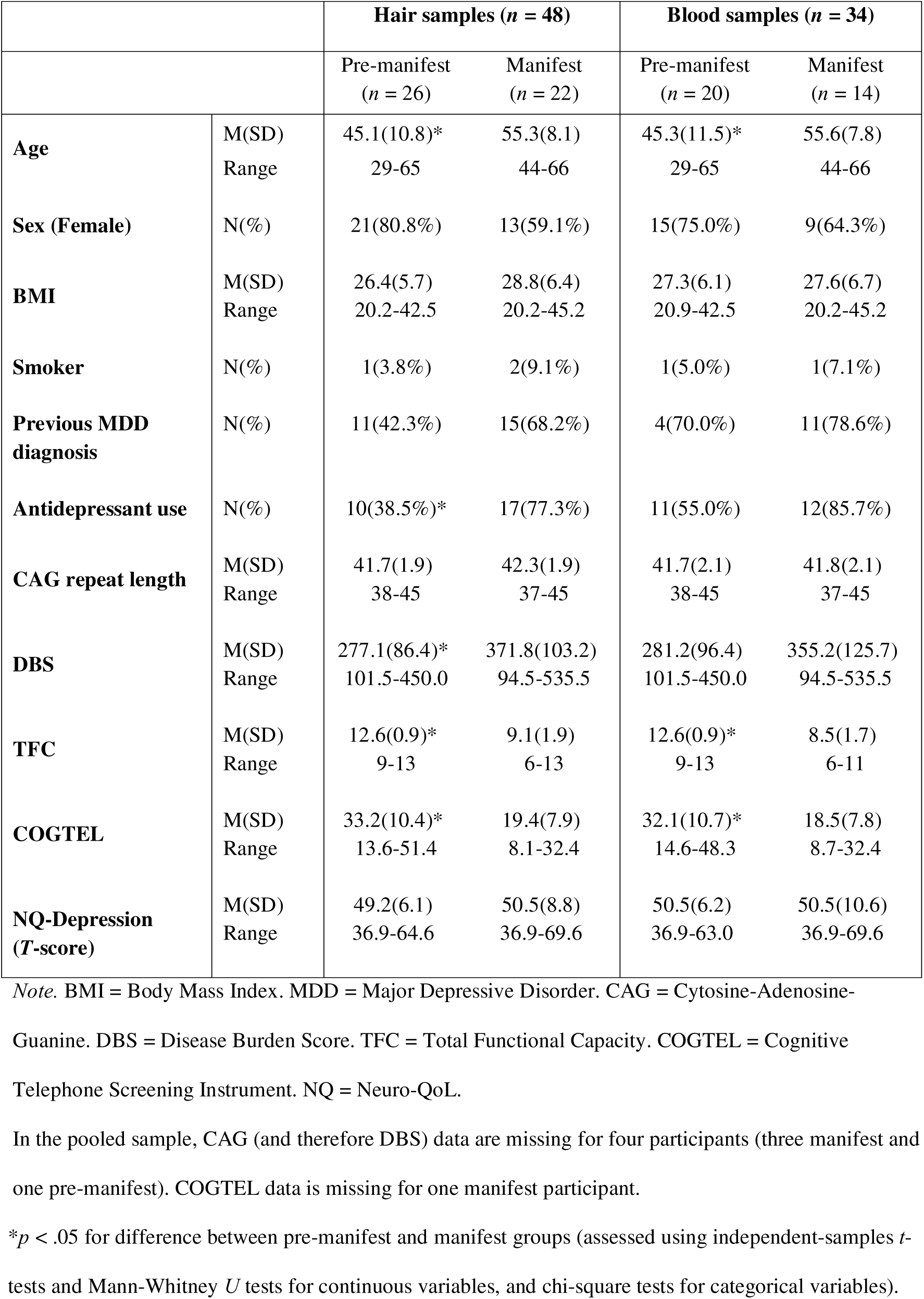
Descriptive Statistics for Hair and Blood Sample Subgroups

## Measures

### Quality of Life in Neurological Disorders (Neuro-QoL) – Depression Questionnaire

Depression severity was self-rated by the participants using the short form of the Neuro-QoL Depression questionnaire [54], which assesses depressive symptoms experienced in the past week. Neuro-QoL Depression is weighted towards the non-somatic symptoms of depression, which makes it suitable for use in neurological populations such as HD. This measure is rated on a Likert scale ranging from 1 to 5, with higher scores indicating more severe depressive symptoms. Raw scores were converted into standardised *T*-scores with a mean of 50 and a standard deviation of 10.

### Cognitive Telephone Screening Instrument (COGTEL)

We used the COGTEL [55] to measure performance-based cognitive functioning. The COGTEL is a telephone-based cognitive screening test which consists of six subtests assessing prospective memory (0-1 point), immediate and delayed verbal memory (0-8 points each), working memory (0-12 points), verbal fluency (unlimited points), and inductive reasoning (0-8 points). Scores from each of the six subtests were combined into a weighted total score.

### Hair Cortisol

We used hair cortisol to measure chronic cortisol production and HPA axis functioning. Hair samples were self-collected by participants from the posterior vertex of the head, which yields the most reliable measures of cortisol [56]. Hair samples were stored in a dark, dry area at room temperature until they were ready to be analysed. Hair cortisol was extracted and quantified by Stratech Scientific APAC Pty Ltd (Sydney, Australia). Cortisol levels were quantified in the three centimetres of hair closest to the scalp, where one centimetre of hair reflects approximately one month of cortisol production [45]. Hair samples were first cleaned and allowed to dry for five days before being bathed and sonicated in an extraction solvent for 24 hours. Samples were then removed from the solvent, dried, and reconstituted in phosphate-buffered saline for analysis. Cortisol was analysed in duplicate using a commercially available ELISA assay (Salimetrics, USA) according to the manufacturer’s instructions. Intra assay variability for cortisol analysis was 4.1% and inter assay variability was 4.6%.

### Inflammatory Cytokines

Based on previous research on peripheral inflammation, depression, and cognition, we selected and quantified four inflammatory cytokines (IL-6, IL-10, IL-1β, TNF-α) in blood plasma. IL-6 is a cytokine with both anti- and (predominantly) pro-inflammatory properties, and stimulates the production of acute phase proteins, haematopoiesis, and immune reactions as part of the inflammatory response [57]. IL-1β and TNF-α are pro-inflammatory cytokines that are secreted as part of the innate immune response and influence neuroendocrine and neurotransmitter activity, as well as inducing a constellation of symptoms known as “sickness behaviours” (e.g., fever, reduced food consumption, and social withdrawal) [58, 59]. IL-10 is an anti-inflammatory cytokine that is involved in balancing the immune response via several immunoregulatory effects, such as inhibiting the production of pro-inflammatory cytokines, and enhancing B-cell proliferation and antibody production [60].

Blood was collected from each participant using EDTA tubes at a Melbourne Pathology Services (MPS) collection centre, and blood samples were delivered to a central lab for processing within four hours of collection. Aliquots of 0.5 mL of plasma were extracted and stored at -70°C until analysis. Cytokines were quantified using four highly sensitive, single-plex Advantage assays on the SIMOA HD-X analyser (Quanterix, Billierica, MA, USA). All samples were tested in duplicate. Except for two IL-1β samples, all samples measured above their lower limit of quantification for each inflammatory marker (IL-6: 0.01 pg/mL; IL-10: 0.021 pg/mL; IL-1β: 0.016 pg/mL; TNF-α: 0.016 pg/mL). For the two IL-1β samples which fell below the lower limit of quantification, values were replaced with the lower limit of detection for IL-1β (0.0055 pg/mL). The average coefficient of variation (CV) for duplicate samples was 4.77% for IL-6, 4.32% for IL-10, 21.71% for IL-1β, and 4.06% for TNF-α.

### Study Procedure

All study components were completed remotely as part of a broader four-week investigation of depression in HD. Participants first completed study screening and the TFC scale via telephone with the first author (HB). On day 1 of the study, participants completed the study consent form and Neuro-QoL Depression questionnaire electronically via a web-based link to a Research Electronic Data Capture (REDCap) system [61]. Participants completed the COGTEL with HB via telephone, approximately one week after baseline. Participants provided a blood sample at a Melbourne Pathology Services (MPS) collection centre on any weekday during the four-week period of testing. Because inflammatory molecules are sensitive to time of day [62], all blood test appointments were scheduled in the morning, as close as possible to the participant’s usual waking time. Participants also self-collected a hair sample at the end of the four-week period of testing, with the assistance of a companion.

Seven participants provided a blood sample following completion of the four-week study, after restrictions related to COVID-19 had eased; these participants re-completed the COGTEL (alternate form) and Neuro-QoL Depression questionnaire on the same day as their blood draw, so that their cytokine data could be correlated with up-to-date clinical data. Blood and hair samples were collected from three additional participants during on-site research visits at Monash University. These three participants completed the Neuro-QoL Depression questionnaire on the same day as biological sample collection, and the COGTEL with HB via telephone within two weeks.

### Data Analysis

Statistical analyses were conducted using SPSS Version 27. Preliminary assumption testing revealed a significant positive skew for all inflammatory cytokines and hair cortisol, which were log-transformed for subsequent statistical analyses. To examine whether hair cortisol and cytokine concentrations differed between pre-manifest and manifest groups, we performed independent-samples *t*-tests, or Mann-Whitney *U* tests for variables whose distributions continued to deviate from normality after log transformations (i.e., IL-1β, TNF-

α, and hair cortisol). Cohen’s *d* was calculated to obtain effect sizes for all between-groups comparisons. We used Spearman’s rank order correlations to examine correlations among all biological measures, and to quantify associations between each biological measure and depression symptom severity, cognitive performance, demographic variables (i.e., age, sex, body mass index (BMI), smoking status, antidepressant use), disease burden, and functional capacity. We also performed non-parametric partial correlation tests to examine the associations of each biological measure with depression severity and cognitive performance after controlling for age, sex, and BMI. We then performed multiple regression analyses to assess the independent associations of each inflammatory cytokine with depression severity and cognitive performance, and to assess if there were significant interactions between each biological measure and disease stage (pre-manifest and manifest) for depression and cognition. Simple slopes analyses were performed for post-hoc characterisation of statistically significant interactions.

## Results

### Cytokine and Cortisol Concentrations Relative to Demographic and Clinical Variables

As depicted in Figure 1, the pre-manifest and manifest groups did not significantly differ for any biological measure: hair cortisol (*p* = .56, *d* = 0.17), IL-6 (*p* = .12, *d* = 0.56), IL-10 (*p* = .06, *d* = 0.70), IL-1β (*p* = .55, *d* = 0.22) or TNF-α (*p* = .27, *d* = 0.40), although medium effect sizes were observed for IL-6 and IL-10.

**Figure 1.**
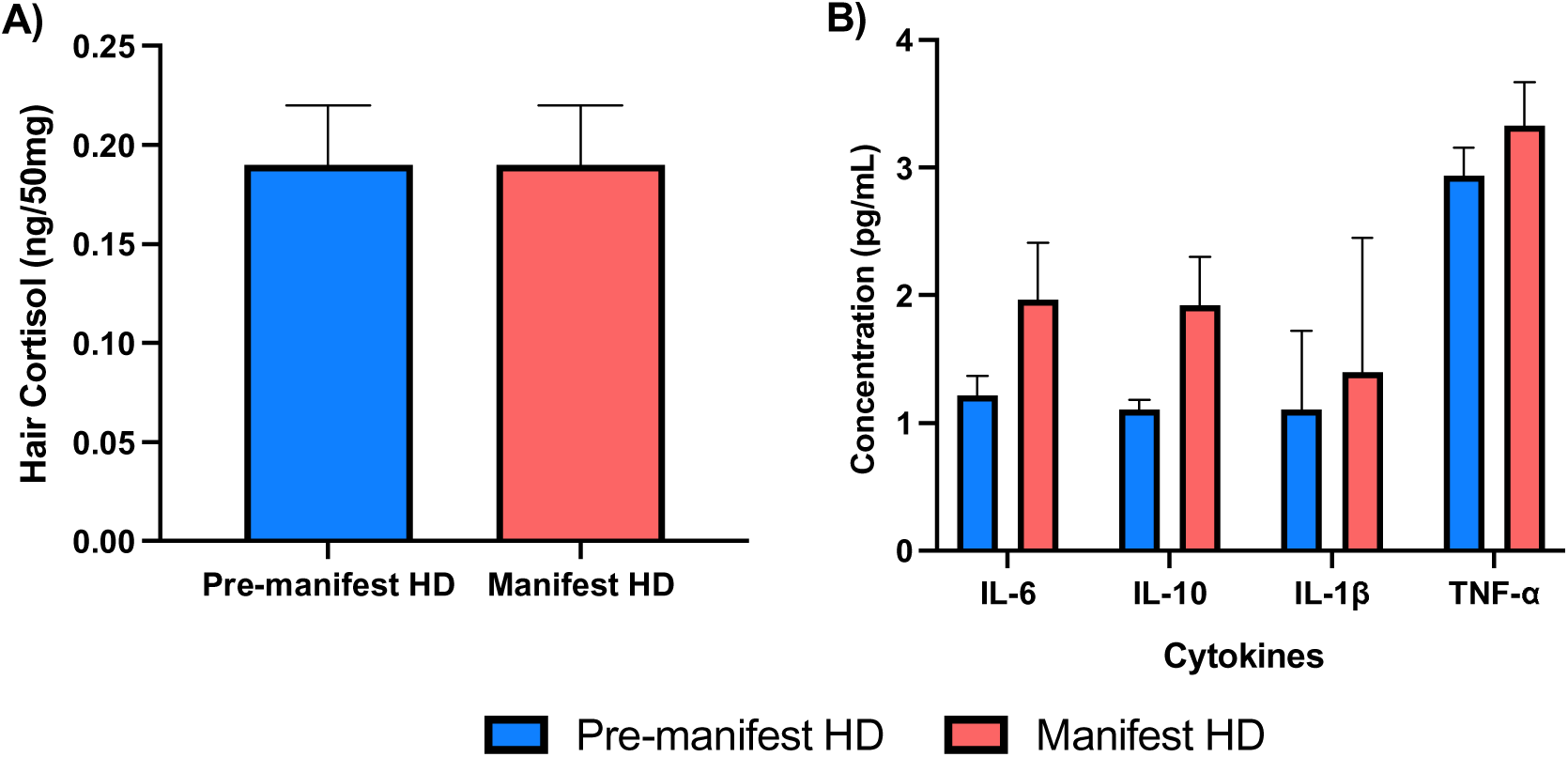
**Hair Cortisol and Inflammatory Cytokine Concentrations in Pre-Manifest and Manifest Groups** *Note.* Raw values are presented for hair cortisol and inflammatory cytokines; however, between-groups comparisons were performed on log-transformed data. Error bars = standard error of the mean.

Higher concentrations of IL-6 significantly correlated with older age, higher BMI, and current use of antidepressants (see Table 2). Moreover, higher concentrations of IL-10 significantly correlated with older age, antidepressant use, and poorer functional capacity, while higher concentrations of TNF-α correlated with older age and antidepressant use. Hair cortisol did not significantly correlate with any demographic or clinical variables.

**Table 2.** Spearman’s Correlations Between Biological Measures and Demographic and Clinical Variables

| | Hair cortisol<sup>a</sup> | | IL-6<sup>b</sup> | | IL-10<sup>b</sup> | | IL-1β<sup>b</sup> | | TNF-α<sup>b</sup> | |
| --- | --- | --- | --- | --- | --- | --- | --- | --- | --- | --- |
| | $r_s$ | $p$ | $r_s$ | $p$ | $r_s$ | $p$ | $r_s$ | $p$ | $r_s$ | $p$ |
| Age | .005 | .98 | **.35** | **.04** | **.39** | **.02** | .11 | .55 | **.48** | **.004** |
| Sex | .003 | .98 | -.10 | .71 | .16 | .37 | -.30 | .09 | .09 | .60 |
| BMI | .06 | .68 | **.47** | **.006** | .22 | .22 | .22 | .21 | .17 | .36 |
| Smoking status | -.18 | .21 | .01 | .94 | .19 | .28 | .17 | .35 | -.04 | .83 |
| Antidepressant use | .03 | .83 | **.47** | **.006** | **.62** | **<.001** | .20 | .25 | **.36** | **.04** |
| DBS | -.10 | .53 | .05 | .80 | .27 | .14 | .09 | .62 | .13 | .48 |
| TFC | -.21 | .16 | -.31 | .08 | **-.44** | **.01** | -.11 | .55 | -.25 | .17 |

*Note.* Correlation tests were performed on log-transformed values of cortisol and cytokine data. $r_s$ =
Spearman's rho. <sup>a</sup> $n = 48$ . <sup>b</sup> $n = 34$ . Significant correlations ( $p < .05$ ) are in bold text.

With respect to correlations among the biological measures, higher levels of IL-6 significantly correlated with higher levels of IL-10 (*r*_s_ = .45, *p* = .008) and TNF-α (*r*_s_ = .35, *p* = .046). Additionally, higher levels of IL-10 significantly correlated with higher levels of TNF-α (*r*_s_ = .43, *p* = .01). No significant intercorrelations were observed for IL-1β or hair cortisol, although we observed trends for correlations between TNF-α and IL-1β (*r*_s_ = .34, *p* = .05), as well as TNF-α and hair cortisol (*r*_s_ = -.35, *p* = .06).

### Associations of Depression and Cognition with Cytokine and Cortisol Concentrations

In unadjusted analyses, higher concentrations of IL-6, IL-10, and TNF-α significantly correlated with higher (more severe) depression scores, and higher concentrations of IL-10 and TNF-α significantly correlated with poorer cognitive performance; medium to large effect sizes were observed for all significant correlations (see Table 3 and Figure 2). Neither hair cortisol nor IL-1β had significant correlations with depression symptoms or cognitive performance. After including age, sex, and BMI as covariates, only the correlations of IL-10 with depression and cognitive performance remained statistically significant, although a trend was noted for the correlation between TNF-α and depression symptoms (see Table 3).

**Figure 2.**
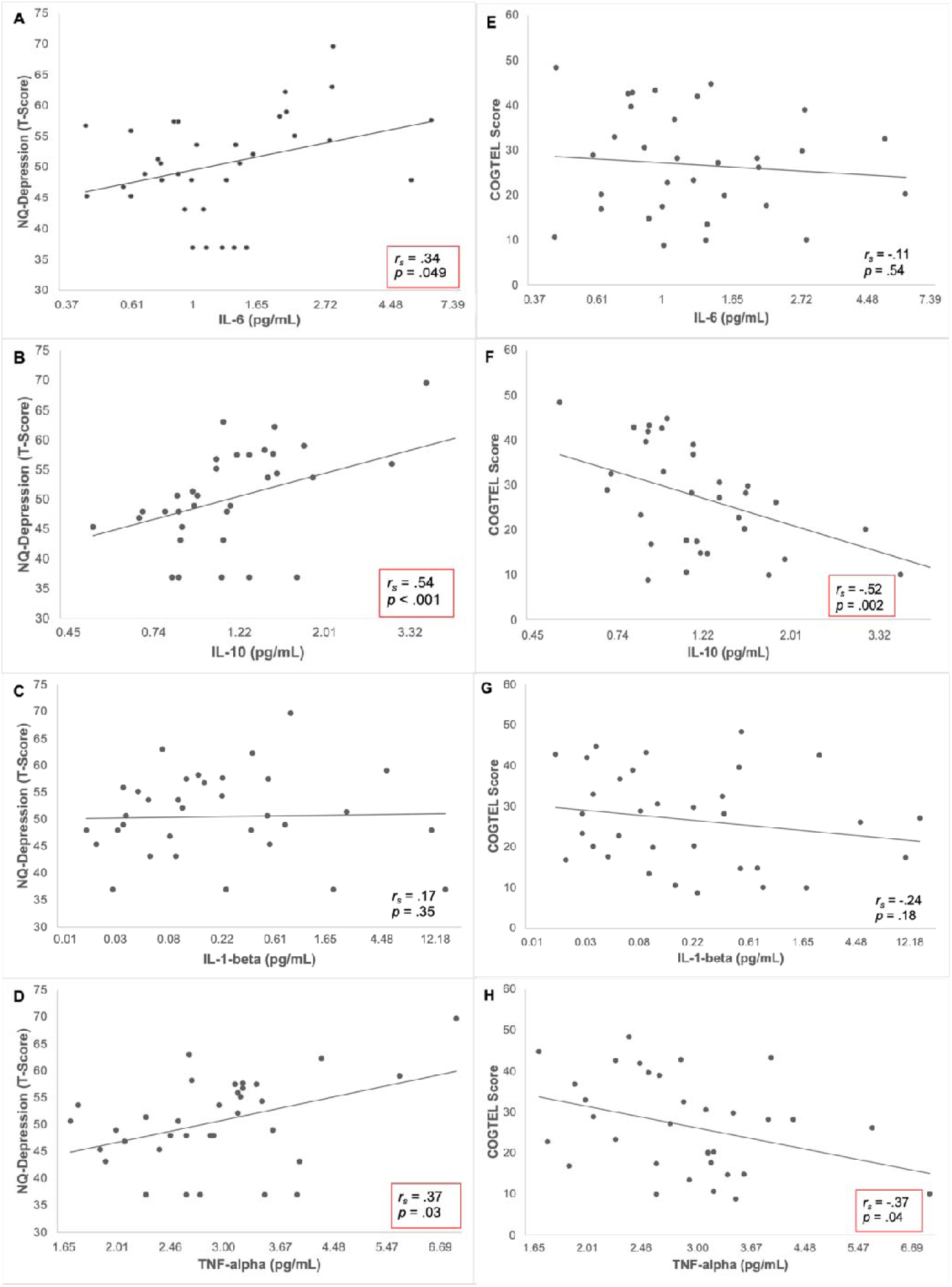
**Associations of Inflammatory Cytokines with Depression Symptoms and Cognitive Performance** *Note.* Scatterplots are presented for relationships of inflammatory cytokines with depression symptoms (left panels; A-D) and cognitive performance (right panels; E-H). Text presented in a red box denotes significant correlations when no covariates were included. Scatterplots present raw values for each variable using a logarithmic scale; all analyses were performed on log-transformed data for each inflammatory cytokine. NQ = Neuro-QoL. *r*_s_ = Spearman’s rho.

**Table 3.** Correlations of Hair Cortisol and Inflammatory Cytokines with Depression Symptom Severity and Cognitive Performance

|  | **Hair cortisol** |  | **IL-6** |  | **IL-10** |  | **IL-1β** |  | **TNF-α** |  |
| --- | --- | --- | --- | --- | --- | --- | --- | --- | --- | --- |
|  | *r<sub>s</sub>* | *p* | *r<sub>s</sub>* | *p* | *r<sub>s</sub>* | *p* | *r<sub>s</sub>* | *p* | *r<sub>s</sub>* | *p* |
| **Unadjusted** |  |  |  |  |  |  |  |  |  |  |
| NQ-Depression | -.01 | .96 | **.34** | **.049** | **.54** | **<.001** | .17 | .35 | **.37** | **.03** |
| COGTEL | .07 | .65 | -.11 | .54 | **-.52** | **.002** | -.24 | .18 | **-.37** | **.04** |
| **Controlling for Age, Sex & BMI** |  |  |  |  |  |  |  |  |  |  |
| NQ-Depression | -.03 | .85 | .26 | .16 | **.53** | **.002** | .17 | .37 | .36 | .05 |
| COGTEL | .10 | .53 | .17 | .37 | **-.41** | **.03** | -.19 | .31 | -.18 | .34 |

*Note.* Correlation tests were performed on log-transformed cortisol and cytokine data. Significant correlations ( $p < .05$ ) are in bold text.

Multiple regression analyses indicated that IL-6, IL-10, TNF-α, and IL-1β together accounted for 30.4% of the variance in depression scores, (*F*(4, 33) = 3.16, *p* = .03), and 29.8% of the variance in cognitive performance (*F*(4, 32) = 2.97, *p* = .04). When included together in the same multiple regression model, none of the cytokines had statistically significant independent associations with depression: IL-6 (*p* = .35), TNF-α (*p* = .27), IL-1β (*p* = .48), although a trend was observed for IL-10 (*p* = .07). Cognitive performance had a significant independent association with IL-10 (β = -.48, *p* = .02), but not with IL-6 (*p* = .49), TNF-α (*p* = .53), or IL-1β (*p* = .58).

### Interactions of Cortisol and Cytokine Concentrations with Disease Stage

As illustrated in Figure 3A, a significant interaction between TNF-α and disease stage was evident for depression symptoms (β = .49, *p* = .02). Specifically, higher concentrations of TNF-α were significantly associated with higher (more severe) depression scores in the manifest group (*r* = .67, *p* = .009), but not in the pre-manifest group (*r* = .14, *p* = .55). Interactions with disease stage were not statistically significant for IL-6 (*p* = .55), IL-10 (*p* = .99), IL-1β (*p* = .33), or hair cortisol (*p* = .24).

**Figure 3.**
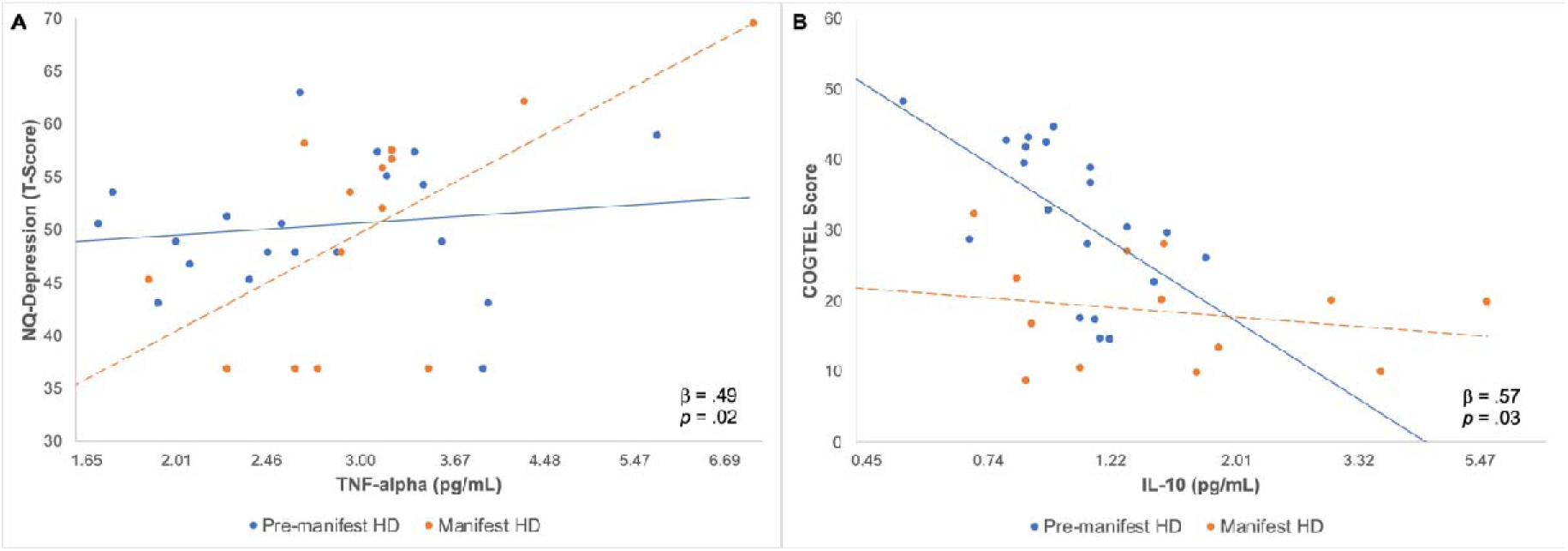
Statistically Significant Interactions Between Biological Measures and Disease Stage for Depression Symptoms and Cognitive Performance *Note.* Scatterplots are presented for the relationship between (A) TNF-α and depression symptoms, and (B) IL-10 and cognitive performance. Scatterplots depict raw values for each variable using a logarithmic scale.

As illustrated in Figure 3B, a significant interaction between disease stage and IL-10 was observed for cognitive performance (β = .57, *p* = .03). Higher concentrations of IL-10 were associated with poorer cognitive performance in the pre-manifest group (*r* = -.63, *p* =

.003), whereas no significant associations were observed between IL-10 and cognitive performance in the manifest group (*r* = -.22, *p* = .47; Figure 3B). Interactions with disease stage were not significant for IL-6 (*p* = .16), IL-1β (*p* = .27), TNF-α (*p* = .57) or hair cortisol (*p* = .88).

## Discussion

This study investigated the associations of systemic inflammation and chronic HPA axis activity with depression symptoms and cognitive functioning in HD. We report that more severe depression symptoms were associated with higher plasma levels of the inflammatory cytokines IL-6, IL-10, and TNF-α, and poorer cognitive performance was associated with higher levels of IL-10 and TNF-α. Conversely, we did not observe any significant associations between hair cortisol concentrations and depression or cognitive performance. We also found that the associations between TNF-α and depression symptoms, and between IL-10 and cognitive performance, differed significantly between pre-manifest and manifest groups. Our findings provide evidence for a relationship between peripheral inflammation and cognitive and psychiatric disturbances in HD, which may have important implications for disease modelling and treatment of HD symptoms.

Our findings indicate that depression symptoms and cognitive disturbances in HD are associated with markers of general immune reactivity (IL-6), pro-inflammatory processes (TNF-α) and anti-inflammatory activity (IL-10). These findings suggest that chronic immune activation may influence the onset, severity, and progression of depression and cognitive difficulties in HD. Our results are in keeping with the large body of research indicating that depression and cognition are associated with elevations in inflammatory cytokines like IL-6, IL-10, and TNF-α in the periphery, both in neurologically healthy populations [21, 63], and in neurodegenerative diseases like Alzheimer’s and Parkinson’s disease [20, 23, 24]. Our results are also partially consistent with a previous study in HD, in which higher levels of IL-6 and IL-1ra were associated with poorer executive functioning [44], although no significant associations were observed between inflammatory cytokines and depression scores.

Interestingly, we found that depression symptoms and poorer cognitive performance had the strongest associations with IL-10, an anti-inflammatory cytokine; these associations remained statistically significant even after controlling for demographic factors and were independent of the effects of other inflammatory cytokines. IL-10 is elevated in many conditions involving CNS pathology, including HD, likely to regulate the production of pro-inflammatory cytokines and thus reduce inflammation [40, 60]. Yet, the elevations previously observed in both pro- and anti-inflammatory cytokines in HD suggest that immune dysregulation persists [36, 38, 40]. In turn, the uncontrolled and prolonged over-expression of inflammatory markers may contribute to depression symptoms and cognitive impairment in HD. Unlike IL-10, the associations of depression and cognitive performance with TNF-α and IL-6 did not remain statistically significant after adjusting for age, sex, and BMI, and should therefore be interpreted with consideration of potential confounding factors. However, the observation of medium effect sizes in unadjusted correlation analyses motivate further investigation and replication of these relationships in larger samples.

We note that IL-1β was not significantly associated with depression symptoms or cognitive performance in our study, despite showing elevations in animal models of HD [64, 65] and being linked to depression and cognitive functioning in non-HD populations [66, 67]. Notably, evidence regarding the associations of IL-1β with depression and cognition is less consistent relative to IL-6 and TNF-α, and significant heterogeneity across previous studies has been reported [21, 50]. The inconsistencies in previous findings may result from difficulties in detecting IL-1β in blood samples (which were also encountered to an extent in this study), and possibly from selective effects of IL-1β in specific subgroups of depressed patients, such as females and people who have experienced early life adversity [68].

Intriguingly, pro-inflammatory activity, represented by TNF-α, was significantly associated with depressive symptoms in the manifest group but not the pre-manifest group, even though TNF-α levels and depression scores were comparable in both groups. This finding reinforces the notion that the aetiology of depression in HD is complex and may vary across disease stages. For example, psychosocial factors and pathophysiological processes other than inflammation may contribute more strongly to depressive symptoms in pre-manifest HD, whereas inflammation may be more relevant to depressive symptomatology in the manifest stage. We also found that anti-inflammatory activity, represented by IL-10, was significantly associated with worse cognitive performance in the pre-manifest group, but not the manifest group. Several studies have shown that inflammatory markers in the CNS and periphery are elevated before the onset of clinical symptoms of HD [33, 34, 36], which suggests that inflammation emerges early in the course of HD. Accordingly, low-grade inflammation may disrupt cognitive functioning in pre-manifest HD, when other pathophysiological changes related to HD are more subtle [7, 16]. In the manifest stage, neurodegeneration and other pathophysiological processes related to HD may become more pronounced, and thus mask or outweigh the effects of inflammation on cognitive functioning.

Several possible mechanisms may underpin the relationship of peripheral inflammation with depression and cognition in HD. Animal studies suggest that peripheral inflammatory cytokines may elicit depression and cognitive changes by crossing the blood brain barrier to influence brain structure and function, or by communicating with the brain via peripheral nerves and endothelial cells of the blood-brain barrier [21, 69]. Alternatively, peripheral inflammatory cytokines may be generated by glial cells in the CNS in response to progressive neurodegeneration and other pathological changes in HD [70] and thus represent a proxy for the actual mechanisms that are causing depression and cognitive impairments in HD. Microglial activation and pro-inflammatory cytokine levels in cerebrospinal fluid have been found to correlate with peripheral pro-inflammatory cytokine levels [36, 71], and both mouse models and humans with HD have previously demonstrated evidence of a more permeable or “leaky” blood brain barrier [72], which supports interactions between immune processes in the CNS and periphery. However, further research is needed to clarify whether peripheral cytokines directly influence depression and cognitive functioning in HD or simply reflect the pathologies that are driving these changes.

Our findings suggest that inflammation is a promising target for the treatment of depression and cognitive symptoms in HD. Pre-clinical studies have previously demonstrated that reducing inflammation improves neuronal, neuropathological, motor, and behavioural abnormalities in mouse models of HD [73–78]. Clinical trials have also yielded promising results, with the LEGATO-HD trial showing that the immunomodulatory drug *laquinimod* is effective in reducing caudate atrophy in HD patients [79], and the SIGNAL-HD trial showing beneficial effects of the drug *pepinemab* on caudate brain atrophy, brain metabolic activity, and an index of global cognitive functioning [80]. Motor functioning and brain atrophy are typically the primary endpoints in HD clinical trials; however, our findings support the inclusion of cognitive functioning and depression as additional outcomes of interest in future trials targeting inflammation in HD. This is particularly salient for future trials in pre-manifest groups, where cognitive and psychiatric symptoms are evident and clinically relevant even in the absence of motor features. Disease stage should also be an important consideration in clinical trials targeting inflammation in HD, as our findings suggest that inflammation may be more relevant to depression and cognitive impairment in specific stages of HD.

Our study suggests that chronic HPA axis functioning may not be strongly associated with depression or cognitive functioning in HD. Only one previous study has assessed hair cortisol concentrations in relation to clinical and disease-related outcomes in pre-manifest HD [43]; like our study, these investigators did not find significant associations between hair cortisol and depression symptoms or any cognitive measures. Moreover, studies in HD which have quantified cortisol levels in bodily fluids have obtained inconsistent findings regarding associations with depression and cognition. For example, two studies in HD have reported associations between salivary cortisol and depression symptoms [29, 30], whereas two other studies found no significant associations between depression severity and plasma or salivary cortisol levels [42, 43]. Similarly, one study has reported that salivary cortisol levels are associated with performance on verbal learning and memory tasks [41], whereas another study found no significant associations between salivary cortisol and cognitive measures [43]. The disparities in findings across studies may be explained by a range of methodological factors, including differences in cortisol collection and quantification, depression and cognitive measures, and sample characteristics across studies. Previous studies also indicate that many of the factors that contribute to depression and cognitive disturbances in HD are unique to the disease [17, 81, 82]. Therefore, unlike neurologically healthy populations, HPA axis functioning may not be strongly implicated in the aetiology of depression and cognitive disturbances in HD.

Several limitations of this study should be noted. Firstly, this study had a relatively small sample size, particularly for the cytokine data, which may have limited statistical power to detect important effects. We have noted several interesting trends in our cytokine data, which did not reach statistical significance but warrant follow-up in larger samples. Further, because this study did not include participants with advanced HD, the relationships of inflammatory cytokines and cortisol with depression and cognition across all disease stages is unknown. This study also did not include a healthy comparison group, which makes it difficult to determine the extent to which the associations observed in this study are unique to HD. Moreover, because of the cross-sectional nature of this study, the directionality of the associations observed between cytokines, depression, and cognition cannot be established with certainty.

In conclusion, this is the first study to show significant associations between systemic inflammation and depression symptoms in pre-manifest and manifest HD. This study also extends previous research showing that peripheral inflammation is linked to poorer cognitive functioning in HD. We did not find significant associations between chronic HPA axis functioning and depression symptoms or cognitive performance in HD. Our findings highlight peripheral inflammation as a possible mechanism underpinning depression and cognitive difficulties in HD. The results of this study also motivate further research on the suitability of inflammatory cytokines as treatment targets for key HD symptoms, and the role of interactions between the immune system and nervous system in the development, exacerbation, and/or progression of HD symptoms.

## Data Availability

All data produced in the present study are available upon reasonable request to the authors.

## Acknowledgements

We thank those who took the time to participate in this study. We also thank Calvary Healthcare Bethlehem and the Neuropsychiatry Team at the Royal Melbourne Hospital for their assistance with recruitment.

## Declarations

### Ethics Approval

The Monash University Human Research Ethics Committee (MUHREC) approved this study (MUHREC IDs: 23043 and 18894). All participants provided written informed consent in accordance with the Declaration of Helsinki.

### Consent

Informed consent was obtained from all individual participants included in the study.

### Author Contribution Statement

All authors contributed to the study conception and design. Material preparation, data collection and analysis were performed by Hiba Bilal. The first draft of the manuscript was written by Hiba Bilal and all authors commented on previous versions of the manuscript. All authors read and approved the final manuscript.

## References

1. McColgan P, Tabrizi SJ (2018) Huntington’s disease: a clinical review. Eur J Neurol 25:24–34. https://doi.org/10.1111/ene.13413

2. Ross CA, Reilmann R, Cardoso F, McCusker EA, Testa CM, Stout JC, Leavitt BR, Pei Z, Landwehrmeyer B, Martinez A, Levey J, Srajer T, Bang J, Tabrizi SJ (2019) Movement Disorder Society Task Force Viewpoint: Huntington’s Disease diagnostic categories. Mov Disord Clin Pract 6:541–546. https://doi.org/10.1002/mdc3.12808

3. Carlozzi NE, Tulsky DS (2013) Identification of health-related quality of life (HRQOL) issues relevant to individuals with Huntington disease. J Health Psychol 18:212–225. https://doi.org/10.1177/1359105312438109

4. Glidden AM, Luebbe EA, Elson MJ, Goldenthal SB, Snyder CW, Zizzi CE, Dorsey ER, Heatwole CR (2020) Patient-reported impact of symptoms in Huntington disease: PRISM-HD. Neurology 94:e2045–e2053. https://doi.org/10.1212/WNL.0000000000008906

5. U.S. Food and Drug Administration (2016) The Voice of the Patient. In: A series of reports from the U.S. Food and Drug Administration’s (FDA’s) Patient-Focused Drug Development Initiative. Center for Drug Evaluation and Research (CDER), pp 1–24

6. Duff K, Paulsen JS, Beglinger LJ, Langbehn DR, Stout JC (2007) Psychiatric symptoms in Huntington’s disease before diagnosis: the PREDICT-HD study. Biol Psychiatry 62:1341–1346. https://doi.org/10.1016/j.biopsych.2006.11.034

7. Tabrizi SJ, Langbehn DR, Leavitt BR, Roos RA, Durr A, Craufurd D, Kennard C, Hicks SL, Fox NC, Scahill RI, Borowsky B, Tobin AJ, Rosas HD, Johnson H, Reilmann R, Landwehrmeyer B, Stout JC, TRACK-HD Investigators (2009) Biological and clinical manifestations of Huntington’s disease in the longitudinal TRACK-HD study: cross-sectional analysis of baseline data. Lancet Neurol 8:791–801. https://doi.org/10.1016/S1474-4422(09)70170-X

8. Stout JC, Paulsen JS, Queller S, Solomon AC, Whitlock KB, Campbell JC, Carlozzi N, Duff K, Beglinger LJ, Langbehn DR, Johnson SA, Biglan KM, Aylward EH (2011) Neurocognitive signs in prodromal Huntington disease. Neuropsychology 25:1–14. https://doi.org/10.1037/a0020937

9. Martinez-Horta S, Perez-Perez J, van Duijn E, Fernandez-Bobadilla R, Carceller M, Pagonabarraga J, Pascual-Sedano B, Campolongo A, Ruiz-Idiago J, Sampedro F, Landwehrmeyer GB, Spanish REGISTRY Investigators of the European Huntington’s Disease Network, Kulisevsky J (2016) Neuropsychiatric symptoms are very common in premanifest and early stage Huntington’s Disease. Parkinsonism Relat Disord 25:58–64. https://doi.org/10.1016/j.parkreldis.2016.02.008

10. Paoli RA, Botturi A, Ciammola A, Silani V, Prunas C, Lucchiari C, Zugno E, Caletti E (2017) Neuropsychiatric burden in Huntington’s disease. Brain Sci 7. https://doi.org/10.3390/brainsci7060067

11. Paulsen JS, Nehl C, Hoth KF, Kanz JE, Benjamin M, Conybeare R, McDowell B, Turner B (2005) Depression and stages of Huntington’s disease. J Neuropsychiatry Clin Neurosci 17:496–502.

12. Thompson JC, Harris J, Sollom AC, Stopford CL, Howard E, Snowden JS, Craufurd D (2012) Longitudinal evaluation of neuropsychiatric symptoms in Huntington’s disease. J Neuropsychiatry Clin Neurosci 24:53–60.

13. Connors MH, Teixeira-Pinto A, Loy CT (2023) Apathy and depression in Huntington’s disease: distinct longitudinal trajectories and clinical correlates. J Neuropsychiatry Clin Neurosci 35:69–76. https://doi.org/10.1176/appi.neuropsych.21070191

14. Epping EA, Mills JA, Beglinger LJ, Fiedorowicz JG, Craufurd D, Smith MM, Groves M, Bijanki KR, Downing N, Williams JK, Long JD, Paulsen JS, Predict-HD Investigators, Coordinators of the Huntington Study Group (2013) Characterization of depression in prodromal Huntington disease in the neurobiological predictors of HD (PREDICT-HD) study. J Psychiatr Res 47:1423–1431. https://doi.org/10.1016/j.jpsychires.2013.05.026

15. Epping EA, Paulsen JS (2011) Depression in the early stages of Huntington disease. Neurodegener Dis Manag 1:407–414. https://doi.org/10.2217/nmt.11.45

16. Tabrizi SJ, Scahill RI, Durr A, Roos RA, Leavitt BR, Jones R, Landwehrmeyer GB, Fox NC, Johnson H, Hicks SL, Kennard C, Craufurd D, Frost C, Langbehn DR, Reilmann R, Stout JC, Track-HD Investigators (2011) Biological and clinical changes in premanifest and early stage Huntington’s disease in the TRACK-HD study: the 12-month longitudinal analysis. Lancet Neurol 10:31–42. https://doi.org/10.1016/S1474-4422(10)70276-3

17. Montoya A, Price BH, Menear M, Lepage M (2006) Brain imaging and cognitive dysfunctions in Huntington’s disease. J Psychiatry Neurosci 31:21–29.

18. Papoutsi M, Labuschagne I, Tabrizi SJ, Stout JC (2014) The cognitive burden in Huntington’s disease: pathology, phenotype, and mechanisms of compensation. Mov Disord 29:673–683. https://doi.org/10.1002/mds.25864

19. Martínez-Horta S, Perez-Perez J, Oltra-Cucarella J, Sampedro F, Horta-Barba A, Puig-Davi A, Pagonabarraga J, Kulisevsky J (2023) Divergent cognitive trajectories in early-stage Huntington’s disease: A three-year longitudinal study. Eur J Neurol. https://doi.org/10.1111/ene.15806

20. Khemka VK, Ganguly A, Bagchi D, Ghosh A, Bir A, Biswas A, Chattopadhyay S, Chakrabarti S (2014) Raised serum proinflammatory cytokines in Alzheimer’s disease with depression. Aging Dis 5:170–176. https://doi.org/10.14336/AD.2014.0500170

21. Kohler CA, Freitas TH, Maes M, de Andrade NQ, Liu CS, Fernandes BS, Stubbs B, Solmi M, Veronese N, Herrmann N, Raison CL, Miller BJ, Lanctot KL, Carvalho AF (2017) Peripheral cytokine and chemokine alterations in depression: a meta-analysis of 82 studies. Acta Psychiatr Scand 135:373–387. https://doi.org/10.1111/acps.12698

22. Lindqvist D, Hall S, Surova Y, Nielsen HM, Janelidze S, Brundin L, Hansson O (2013) Cerebrospinal fluid inflammatory markers in Parkinson’s disease--associations with depression, fatigue, and cognitive impairment. Brain Behav Immun 33:183–189. https://doi.org/10.1016/j.bbi.2013.07.007

23. Lindqvist D, Kaufman E, Brundin L, Hall S, Surova Y, Hansson O (2012) Non-motor symptoms in patients with Parkinson’s disease - correlations with inflammatory cytokines in serum. PLoS One 7:e47387. https://doi.org/10.1371/journal.pone.0047387

24. Menza M, DeFronzo Dobkin R, Marin H, Mark MH, Gara M, Bienfait K, Dicke A, Kusnekov A (2010) The role of inflammatory cytokines in cognition and other non-motor symptoms of Parkinson’s disease. Psychosomatics 51:474–479. https://doi.org/10.1016/s0033-3182(10)70739-8

25. Nikkheslat N, Pariante CM, Zunszain PA (2018) Neuroendocrine Abnormalities in Major Depression: An Insight Into Glucocorticoids, Cytokines, and the Kynurenine Pathway. In: Inflammation and Immunity in Depression. pp 45–60

26. Pariante CM (2017) Why are depressed patients inflamed? A reflection on 20 years of research on depression, glucocorticoid resistance and inflammation. Eur Neuropsychopharmacol 27:554–559. https://doi.org/10.1016/j.euroneuro.2017.04.001

27. Piantella S, O’Brien WT, Hale MW, Maruff P, McDonald SJ, Wright BJ (2022) Within subject rise in serum TNFα to IL-10 ratio is associated with poorer attention, decision-making and working memory in jockeys. Comprehensive Psychoneuroendocrinology 10:100131. https://doi.org/https://doi.org/10.1016/j.cpnec.2022.100131

28. Aziz NA, Pijl H, Frolich M, van der Graaf AW, Roelfsema F, Roos RA (2009) Increased hypothalamic-pituitary-adrenal axis activity in Huntington’s disease. J Clin Endocrinol Metab 94:1223–1228. https://doi.org/10.1210/jc.2008-2543

29. Hubers AA, van der Mast RC, Pereira AM, Roos RA, Veen LJ, Cobbaert CM, van Duijn E, Giltay EJ (2015) Hypothalamic-pituitary-adrenal axis functioning in Huntington’s disease and its association with depressive symptoms and suicidality. J Neuroendocrinol 27:234–244. https://doi.org/10.1111/jne.12255

30. Shirbin CA, Chua P, Churchyard A, Lowndes G, Hannan AJ, Pang TY, Chiu E, Stout JC (2013) Cortisol and depression in pre-diagnosed and early stage Huntington’s disease. Psychoneuroendocrinology 38:2439–2447. https://doi.org/10.1016/j.psyneuen.2012.10.020

31. van Duijn E, Selis MA, Giltay EJ, Zitman FG, Roos RA, van Pelt H, van der Mast RC (2010) Hypothalamic-pituitary-adrenal axis functioning in Huntington’s disease mutation carriers compared with mutation-negative first-degree controls. Brain Res Bull 83:232–237. https://doi.org/10.1016/j.brainresbull.2010.08.006

32. Pavese N, Gerhard A, Tai YF, Ho AK, Turkheimer F, Barker RA, Brooks DJ, Piccini P (2006) Microglial activation correlates with severity in Huntington disease: a clinical and PET study. Neurology 66:1638–1643. https://doi.org/10.1212/01.wnl.0000222734.56412.17

33. Politis M, Pavese N, Tai YF, Kiferle L, Mason SL, Brooks DJ, Tabrizi SJ, Barker RA, Piccini P (2011) Microglial activation in regions related to cognitive function predicts disease onset in Huntington’s disease: a multimodal imaging study. Hum Brain Mapp 32:258–270. https://doi.org/10.1002/hbm.21008

34. Tai YF, Pavese N, Gerhard A, Tabrizi SJ, Barker RA, Brooks DJ, Piccini P (2007) Microglial activation in presymptomatic Huntington’s disease gene carriers. Brain 130:1759–1766. https://doi.org/10.1093/brain/awm044

35. Sapp E, Kegel KB, Aronin N, Hashikawa T, Uchiyama Y, Tohyama K, Bhide PG, Vonsattel JP, DiFiglia M (2001) Early and progressive accumulation of reactive microglia in the Huntington disease brain. J Neuropathol Exp Neurol 60:161–172. https://doi.org/10.1093/jnen/60.2.161

36. Bjorkqvist M, Wild EJ, Thiele J, Silvestroni A, Andre R, Lahiri N, Raibon E, Lee RV, Benn CL, Soulet D, Magnusson A, Woodman B, Landles C, Pouladi MA, Hayden MR, Khalili-Shirazi A, Lowdell MW, Brundin P, Bates GP, Leavitt BR, Moller T, Tabrizi SJ (2008) A novel pathogenic pathway of immune activation detectable before clinical onset in Huntington’s disease. J Exp Med 205:1869–1877. https://doi.org/10.1084/jem.20080178

37. Dalrymple A, Wild EJ, Joubert R, Sathasivam K, Björkqvist M, Petersén A, Jackson GS, Isaacs JD, Kristiansen M, Bates GP, Leavitt BR, Keir G, Ward M, Tabrizi SJ (2007) Proteomic profiling of plasma in Huntington’s disease reveals neuroinflammatory activation and biomarker candidates. J Proteome Res 6:2833–2840. https://doi.org/10.1021/pr0700753

38. Chang KH, Wu YR, Chen YC, Chen CM (2015) Plasma inflammatory biomarkers for Huntington’s disease patients and mouse model. Brain Behav Immun 44:121–127. https://doi.org/10.1016/j.bbi.2014.09.011

39. Sanchez-Lopez F, Tasset I, Aguera E, Feijoo M, Fernandez-Bolanos R, Sanchez FM, Ruiz MC, Cruz AH, Gascon F, Tunez I (2012) Oxidative stress and inflammation biomarkers in the blood of patients with Huntington’s disease. Neurol Res 34:721–724. https://doi.org/10.1179/1743132812Y.0000000073

40. Silvestroni A, Faull RL, Strand AD, Moller T (2009) Distinct neuroinflammatory profile in post-mortem human Huntington’s disease. Neuroreport 20:1098–1103. https://doi.org/10.1097/WNR.0b013e32832e34ee

41. Shirbin CA, Chua P, Churchyard A, Hannan AJ, Lowndes G, Stout JC (2013) The relationship between cortisol and verbal memory in the early stages of Huntington’s disease. J Neurol 260:891–902. https://doi.org/10.1007/s00415-012-6732-y

42. Aziz NA, Anguelova GV, Marinus J, Lammers GJ, Roos RA (2010) Sleep and circadian rhythm alterations correlate with depression and cognitive impairment in Huntington’s disease. Parkinsonism Relat Disord 16:345–350. https://doi.org/10.1016/j.parkreldis.2010.02.009

43. Cruickshank T, Porter T, Laws SM, Ziman M, Bartlett DM (2021) Hair and salivary cortisol and their relationship with lifestyle, mood and cognitive outcomes in premanifest Huntington’s disease. Sci Rep 11:5464. https://doi.org/10.1038/s41598-021-84726-4

44. Bouwens JA, van Duijn E, Cobbaert CM, Roos RA, van der Mast RC, Giltay EJ (2016) Plasma cytokine levels in relation to neuropsychiatric symptoms and cognitive dysfunction in Huntington’s disease. J Huntingtons Dis 5:369–377. https://doi.org/10.3233/JHD-160213

45. Russell E, Koren G, Rieder M, Van Uum S (2012) Hair cortisol as a biological marker of chronic stress: current status, future directions and unanswered questions. Psychoneuroendocrinology 37:589–601. https://doi.org/10.1016/j.psyneuen.2011.09.009

46. Feeney JC, O’Halloran AM, Kenny RA (2018) The association between hair cortisol, hair cortisone, and cognitive function in a population-based cohort of older adults: Results from the Irish Longitudinal Study on Ageing. J Gerontol 75:257–265. https://doi.org/10.1093/gerona/gly258

47. Pochigaeva K, Druzhkova T, Yakovlev A, Onufriev M, Grishkina M, Chepelev A, Guekht A, Gulyaeva N (2017) Hair cortisol as a marker of hypothalamic-pituitary-adrenal Axis activity in female patients with major depressive disorder. Metab Brain Dis 32:577–583. https://doi.org/10.1007/s11011-017-9952-0

48. Pulopulos MM, Hidalgo V, Almela M, Puig-Perez S, Villada C, Salvador A (2014) Hair cortisol and cognitive performance in healthy older people. Psychoneuroendocrinology 44:100–111. https://doi.org/10.1016/j.psyneuen.2014.03.002

49. Steudte-Schmiedgen S, Wichmann S, Stalder T, Hilbert K, Muehlhan M, Lueken U, Beesdo-Baum K (2017) Hair cortisol concentrations and cortisol stress reactivity in generalized anxiety disorder, major depression and their comorbidity. J Psychiatr Res 84:184–190. https://doi.org/10.1016/j.jpsychires.2016.09.024

50. Fard MT, Savage KM, Stough CK (2022) Peripheral inflammation marker relationships to cognition in healthy older adults – A systematic review. Psychoneuroendocrinology 144:105870. https://doi.org/https://doi.org/10.1016/j.psyneuen.2022.105870

51. Kay C, Collins JA, Miedzybrodzka Z, Madore SJ, Gordon ES, Gerry N, Davidson M, Slama RA, Hayden MR (2016) Huntington disease reduced penetrance alleles occur at high frequency in the general population. Neurology 87:282–288. https://doi.org/10.1212/wnl.0000000000002858

52. Huntington Study Group (1996) Unified Huntington’s Disease Rating Scale: reliability and consistency. Mov Disord 11:136–142. https://doi.org/10.1002/mds.870110204

53. Penney JB, Vonsattel J-P, Macdonald ME, Gusella JF, Myers RH (1997) CAG repeat number governs the development rate of pathology in Huntington’s disease. Annals of Neurology 41:689–692. https://doi.org/https://doi.org/10.1002/ana.410410521

54. National Institute of Neurological Disorders and Stroke (2015) User Manual for the Quality of Life in Neurological Disorders (Neuro-QoL) Measures, Version 2.0. In:

55. Kliegel M, Martin M, Jager T (2007) Development and validation of the Cognitive Telephone Screening Instrument (COGTEL) for the assessment of cognitive function across adulthood. J Psychol 141:147–170. https://doi.org/10.3200/JRLP.141.2.147-172.

56. Greff MJE, Levine JM, Abuzgaia AM, Elzagallaai AA, Rieder MJ, van Uum SHM (2019) Hair cortisol analysis: An update on methodological considerations and clinical applications. Clin Biochem 63:1–9. https://doi.org/10.1016/j.clinbiochem.2018.09.010

57. Tanaka T, Narazaki M, Kishimoto T (2014) IL-6 in inflammation, immunity, and disease. Cold Spring Harb Perspect Biol 6:a016295. https://doi.org/10.1101/cshperspect.a016295

58. Brebner K, Hayley S, Zacharko R, Merali Z, Anisman H (2000) Synergistic effects of interleukin-1β, interleukin-6, and tumor necrosis factor-α: Central monoamine, corticosterone, and behavioral variations. Neuropsychopharmacology 22:566–580. https://doi.org/10.1016/S0893-133X(99)00166-9

59. Dantzer R (2009) Cytokine, sickness behavior, and depression. Immunol Allergy Clin North Am 29:247–264. https://doi.org/10.1016/j.iac.2009.02.002

60. Porro C, Cianciulli A, Panaro MA (2020) The regulatory role of IL-10 in neurodegenerative diseases. Biomolecules 10. https://doi.org/10.3390/biom10071017

61. Harris PA, Taylor R, Thielke R, Payne J, Gonzalez N, Conde JG (2009) Research electronic data capture (REDCap)--a metadata-driven methodology and workflow process for providing translational research informatics support. J Biomed Inform 42:377–381. https://doi.org/10.1016/j.jbi.2008.08.010

62. Zhou X, Fragala MS, McElhaney JE, Kuchel GA (2010) Conceptual and methodological issues relevant to cytokine and inflammatory marker measurements in clinical research. Curr Opin Clin Nutr Metab Care 13:541–547. https://doi.org/10.1097/MCO

63. Marsland AL, Gianaros PJ, Kuan DC, Sheu LK, Krajina K, Manuck SB (2015) Brain morphology links systemic inflammation to cognitive function in midlife adults. Brain Behav Immun 48:195–204. https://doi.org/10.1016/j.bbi.2015.03.015

64. Pido-Lopez J, Andre R, Benjamin AC, Ali N, Farag S, Tabrizi SJ, Bates GP (2018) In vivo neutralization of the protagonist role of macrophages during the chronic inflammatory stage of Huntington’s disease. Sci Rep 8:11447. https://doi.org/10.1038/s41598-018-29792-x

65. Valekova I, Jarkovska K, Kotrcova E, Bucci J, Ellederova Z, Juhas S, Motlik J, Gadher SJ, Kovarova H (2016) Revelation of the IFNalpha, IL-10, IL-8 and IL-1beta as promising biomarkers reflecting immuno-pathological mechanisms in porcine Huntington’s disease model. J Neuroimmunol 293:71–81. https://doi.org/10.1016/j.jneuroim.2016.02.012

66. Leo R, Di Lorenzo G, Tesauro M, Razzini C, Forleo GB, Chiricolo G, Cola C, Zanasi M, Troisi A, Siracusano A, Lauro R, Romeo F (2006) Association between enhanced soluble CD40 ligand and proinflammatory and prothrombotic states in major depressive disorder: pilot observations on the effects of selective serotonin reuptake inhibitor therapy. J Clin Psychiatry 67:1760–1766. https://doi.org/10.4088/jcp.v67n1114

67. Simpson EE, Hodkinson CF, Maylor EA, McCormack JM, Rae G, Strain S, Alexander HD, Wallace JM (2013) Intracellular cytokine production and cognition in healthy older adults. Psychoneuroendocrinology 38:2196–2208. https://doi.org/10.1016/j.psyneuen.2013.04.007

68. McQuaid RJ, Gabrys RL, McInnis OA, Anisman H, Matheson K (2019) understanding the relation between early-life adversity and depression symptoms: The moderating role of sex and an interleukin-1beta gene variant. Front Psychiatry 10:151. https://doi.org/10.3389/fpsyt.2019.00151

69. Yirmiya R, Goshen I (2011) Immune modulation of learning, memory, neural plasticity and neurogenesis. Brain Behav Immun 25:181–213. https://doi.org/10.1016/j.bbi.2010.10.015

70. Crotti A, Glass CK (2015) The choreography of neuroinflammation in Huntington’s disease. Trends Immunol 36:364–373. https://doi.org/10.1016/j.it.2015.04.007

71. Politis M, Lahiri N, Niccolini F, Su P, Wu K, Giannetti P, Scahill RI, Turkheimer FE, Tabrizi SJ, Piccini P (2015) Increased central microglial activation associated with peripheral cytokine levels in premanifest Huntington’s disease gene carriers. Neurobiol Dis 83:115–121. https://doi.org/10.1016/j.nbd.2015.08.011

72. Drouin-Ouellet J, Sawiak SJ, Cisbani G, Lagace M, Kuan WL, Saint-Pierre M, Dury RJ, Alata W, St-Amour I, Mason SL, Calon F, Lacroix S, Gowland PA, Francis ST, Barker RA, Cicchetti F (2015) Cerebrovascular and blood-brain barrier impairments in Huntington’s disease: Potential implications for its pathophysiology. Ann Neurol 78:160–177. https://doi.org/10.1002/ana.24406

73. Bouchard J, Truong J, Bouchard K, Dunkelberger D, Desrayaud S, Moussaoui S, Tabrizi SJ, Stella N, Muchowski PJ (2012) Cannabinoid receptor 2 signaling in peripheral immune cells modulates disease onset and severity in mouse models of Huntington’s disease. J Neurosci 32:18259–18268. https://doi.org/10.1523/JNEUROSCI.4008-12.2012

74. Garcia-Miralles M, Hong X, Tan LJ, Caron NS, Huang Y, To XV, Lin RY, Franciosi S, Papapetropoulos S, Hayardeny L, Hayden MR, Chuang KH, Pouladi MA (2016) Laquinimod rescues striatal, cortical and white matter pathology and results in modest behavioural improvements in the YAC128 model of Huntington disease. Sci Rep 6:31652. https://doi.org/10.1038/srep31652

75. Hsiao HY, Chen YC, Chen HM, Tu PH, Chern Y (2013) A critical role of astrocyte-mediated nuclear factor-kappaB-dependent inflammation in Huntington’s disease. Hum Mol Genet 22:1826–1842. https://doi.org/10.1093/hmg/ddt036

76. Palazuelos J, Aguado T, Pazos MR, Julien B, Carrasco C, Resel E, Sagredo O, Benito C, Romero J, Azcoitia I, Fernandez-Ruiz J, Guzman M, Galve-Roperh I (2009) Microglial CB2 cannabinoid receptors are neuroprotective in Huntington’s disease excitotoxicity. Brain 132:3152–3164. https://doi.org/10.1093/brain/awp239

77. Sagredo O, Gonzalez S, Aroyo I, Pazos MR, Benito C, Lastres-Becker I, Romero JP, Tolon RM, Mechoulam R, Brouillet E, Romero J, Fernandez-Ruiz J (2009) Cannabinoid CB2 receptor agonists protect the striatum against malonate toxicity: relevance for Huntington’s disease. Glia 57:1154–1167. https://doi.org/10.1002/glia.20838

78. Southwell AL, Franciosi S, Villanueva EB, Xie Y, Winter LA, Veeraraghavan J, Jonason A, Felczak B, Zhang W, Kovalik V, Waltl S, Hall G, Pouladi MA, Smith ES, Bowers WJ, Zauderer M, Hayden MR (2015) Anti-semaphorin 4D immunotherapy ameliorates neuropathology and some cognitive impairment in the YAC128 mouse model of Huntington disease. Neurobiol Dis 76:46–56. https://doi.org/10.1016/j.nbd.2015.01.002

79. Reilmann R, Gordon MF, Anderson KE, Feigin A, Tabrizi SJ, Leavitt BR, Stout JC, Piccini P, Borowsky B, Rynkowski G, et al. (2019) The efficacy and safety results of laquinimod as a treatment for Huntington disease (LEGATO-HD). Neurology 92.

80. Feigin A, Evans EE, Fisher TL, Leonard JE, Smith ES, Reader A, Mishra V, Manber R, Walters KA, Kowarski L, Oakes D, Siemers E, Kieburtz KD, Zauderer M, Huntington Study Group SIGNAL Investigators (2022) Pepinemab antibody blockade of SEMA4D in early Huntington’s disease: a randomized, placebo-controlled, phase 2 trial. Nat Med 28:2183–2193. https://doi.org/10.1038/s41591-022-01919-8

81. Bilal H, Warren N, Dahanayake P, Kelso W, Farrand S, Stout J (2022) The lived experiences of depression in Huntington’s disease: A qualitative study. J Huntingtons Dis 11:1–15. https://doi.org/10.3233/JHD-220537

82. Gubert C, Renoir T, Hannan AJ (2020) Why Woody got the blues: The neurobiology of depression in Huntington’s disease. Neurobiol Dis 142:104958. https://doi.org/10.1016/j.nbd.2020.104958

